# Integration of clinical characteristics, lab tests and a deep learning CT scan analysis to predict severity of hospitalized COVID-19 patients

**DOI:** 10.1101/2020.05.14.20101972

**Authors:** Nathalie Lassau, Samy Ammari, Emilie Chouzenoux, Hugo Gortais, Paul Herent, Matthieu Devilder, Samer Soliman, Olivier Meyrignac, Marie-Pauline Talabard, Jean-Philippe Lamarque, Remy Dubois, Nicolas Loiseau, Paul Trichelair, Etienne Bendjebbar, Gabriel Garcia, Corinne Balleyguier, Mansouria Merad, Annabelle Stoclin, Simon Jegou, Franck Griscelli, Nicolas Tetelboum, Yingping Li, Sagar Verma, Matthieu Terris, Tasnim Dardouri, Kavya Gupta, Ana Neacsu, Frank Chemouni, Meriem Sefta, Paul Jehanno, Imad Bousaid, Yannick Boursin, Emmanuel Planchet, Mikael Azoulay, Jocelyn Dachary, Fabien Brulport, Adrian Gonzalez, Olivier Dehaene, Jean-Baptiste Schiratti, Kathryn Schutte, Jean-Christophe Pesquet, Hugues Talbot, Elodie Pronier, Gilles Wainrib, Thomas Clozel, Fabrice Barlesi, Marie-France Bellin, Michael G. B. Blum

## Abstract

The SARS-COV-2 pandemic has put pressure on Intensive Care Units, so that identifying predictors of disease severity is a priority. We collected 58 clinical and biological variables, chest CT scan data (506,341 images), and radiology reports from 1,003 coronavirus-infected patients from two French hospitals. We trained a deep learning model based on CT scans to predict severity; this model was more discriminative than a radiologist quantification of disease extent. We showed that neural network analysis of CT-scan brings unique prognosis information, although it is correlated with other markers of severity (oxygenation, LDH, and CRP). To provide a multimodal severity score, we developed *AI-severity* that includes 5 clinical and biological variables (age, sex, oxygenation, urea, platelet) as well as the CT deep learning model. When comparing *AI-severity* with 11 existing scores for severity, we find significantly improved prognosis performance; *AI-severity* can therefore rapidly become a reference scoring approach.

## Introduction

Hospitalized COVID-19 patients are likely to develop severe outcomes requiring mechanical ventilation or high-flow oxygenation. Among hospitalized patients, 14 to 30% will require admission to an ICU, 12 to 33% will require mechanical ventilation, and 20% to 33% will die^1–4^. Detection at admission of patients at risk of severe outcomes is important to deliver proper care and to optimize use of limited intensive care unit (ICU) ressources^5^.

Identification of hospitalized COVID-19 patients at risk for severe deterioration can be done using risk scores that combine several factors including age, sex, and comorbidities (CALL, COVID-GRAM, 4C Mortality Score)^6–12^. Some risk scores also include additional markers of severity such as the dyspnea symptom, clinical examination variables such as low oxygen saturation and elevated respiratory rate, as well as biological factors reflecting multi-organ failures such as elevated Lactate dehydrogenase (LDH) values^8,10,13–15^.

Beyond clinical and biological variables, computerized tomography (CT) scans also contain prognostic information, as the degree of pulmonary inflammation is associated with clinical symptoms, and the amount of lung abnormality is associated with severe evolution^16–20^. CT-scans can be acquired at admission to diagnose COVID-19 when RT-PCR results are negative^21^. However, the extent to which CT scans at patient admission add prognostic information beyond what can be inferred from clinical and biological data is unresolved.

The objective of this study was to integrate clinical, biological and radiological data to predict the outcome of hospitalized patients. By processing CT-scan images with a deep learning model and by using a radiologist report that contains a semi-quantitative description of CT-scans, we evaluated the additional amount of information brought by CT-scans.

## Results

A total of 1,003 patients from Kremlin-Bicêtre (KB, Paris, France) and Gustave Roussy (IGR, Villejuif, France) were enrolled in the study. Clinical, biological, and CT scan images and reports were collected at hospital admission. There were 931 patients for which clinical, biological and CT-scan data were available (Supp Fig 1). A total of 506,341 images were analyzed for the 980 patients with CT-scans (average of 517 slices per scan). Radiologists annotated 17,873 images from 329 CT-scans. Summary statistics for the clinical, biological, and CT scan data are provided in Table 1

**Table 1:**
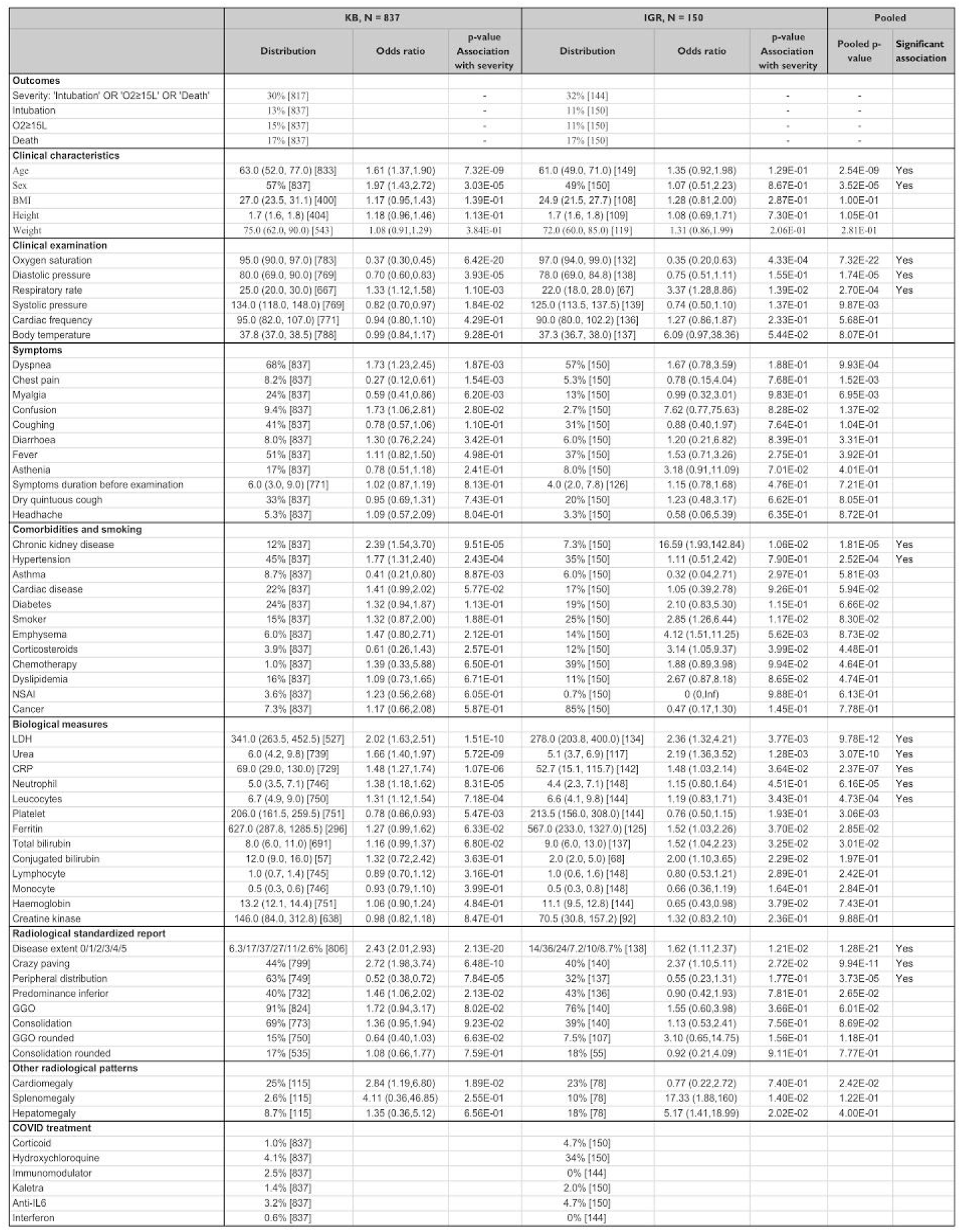
Population description for the KB and IGR hospitals and association between variables measured at admission and severity. Among the 1,003 patients of the study, biological and clinical variables were available for 987 individuals. Categorical variables are expressed as percentages [available]. Continuous variables are shown as median (IQR) [available]. Associations with severity are reported with p-values for each center and p-values were combined with Stouffer’s method. The column entitled “Significant association” indicates that the variable is significantly associated with severity after Bonferroni adjustment to account for multiple testing across 58 variables (treatments are excluded). For continuous variables, odds ratios are computed for an increase of one standard deviation of the continuous variable.

### Variables associated with severity

We first evaluated how clinical and biological variables measured at admission were associated with future severe progression, which we defined as an oxygen flow rate of 15 L/min or higher and/or the need for mechanical ventilation and/or patient death^22^. This definition of severe progression corresponds to a score of 5 or more according to the World Health Organization evaluation of severity on a 1 to 10 scale. We computed the severity odds ratios for each individual variable, and at each hospital center (Table 1). When combining association results from the two centers, we found 12 variables significantly associated with severity (P <0.05/58 to account for testing 58 variables, Table 1): age, sex, oxygen saturation, diastolic pressure, respiratory rate, chronic kidney disease, hypertension, LDH, and urea, CRP, polynuclear neutrophil, and leukocytes.

We then assessed the predictive value of features from admission radiology reports. These reports contain semi-quantitative evaluations of the extent of disease which values range from 0 to 5, as well as a presence/absence coding of several types of lung lesions in COVID-19 patients. We found three significant associated features (P <0.05/58): extent of disease, and presence of crazy paving lesions, which are both associated with greater severity, and presence of a peripheral distribution of lesions, which is associated with lesser severity.

### A neural network model to predict severity based on CT scans

To capture CT-scan prognosis information from images, we considered a weakly supervised approach with no radiologist-provided annotations (Supp Fig 2)^23^. A deep learning model was trained to predict severe progression based on a CT-scan image. The neural network was trained on a development cohort consisting of 646 patients from Kremlin-Bicêtre Hospital (KB). It was evaluated on 150 KB patients, which were left-over from the development cohort, and it was further evaluated using a validation cohort consisting of 135 patients from Institut Gustave Roussy hospital (IGR). The discriminative ability of the neural network was of AUC = 0.76 (0.67,0.85) for the 150 left-over KB patients, which were not used to train the network, and of AUC = 0.75 (0.65,0.84) for the validation IGR dataset. As a point of comparison, the AUC obtained with the radiologist evaluation of disease extent is of 0.73 (0.64-0.82) for the 150 KB patients of the development cohort and of 0.66 (0.56-0.76) for the validation IGR cohort, and the difference between the two AUC values was significant for the validation IGR cohort only (P≤0.05).

### Interpretability analysis of the neural network model

To apprehend the information present within the CT scans that is captured by the weakly supervised neural network model, we evaluated to what extent the features (internal representation) extracted by the neural network can predict clinical and radiological variables. To this end, we trained a new logistic regression with the extracted features as input, and some clinical and radiological variables as output. AUC on the 150 leftover patients of the KB development cohort was 0.93 (0.88,0.97) for disease extent (threshold >2), 0.78 (0.70,0.85) for crazy paving, 0.64 (0.53,0.74) for condensation and 0.80 (0.65,0.94) for Ground Glass Opacity (Supp Table 1). It was also possible to relate internal representations of the neural networks to clinical variables. We obtained an AUC of 0.88 (0.82,0.94) for predicting an age strictly larger than 60 year-old, an AUC of 0.93 (0.89,0.97) for sex, and of 0.76 (0.68,0.84) for predicting an oxygen saturation larger than 90%. As a comparison, a logistic regression trained on the variables from the radiology report obtained only AUC scores of 0.70 (0.61,0.78) for age, 0.57 (0.48, 0.67) for sex and of 0.68 (0.58, 0.77) for oxygen saturation, and differences of AUC were significant (P<0.05). Simply put, this analysis shows that the internal representation of the neural network captures clinical features from the lung CTs, such as sex or age, on top of the known COVID-19 radiology features.

### A multimodal prognostic models for severity

To add information from lab tests and chest characteristics to the CT-scan information, we constructed the *AI-severity* score. We used a greedy search approach to include optimal clinical and biological variables (see Methods). In addition to the CT deep learning variable, the variables included in *AI-severity* are age, sex, oxygen saturation, urea, and platelet counts. Coefficients and transformations required to compute the 6-variable *AI-severity* score are available in Supp Table 2. Coefficients required to compute *AI-severity* were learned using the WHO-defined high severity outcome of “oxygen flow rate of 15 L/min or higher, or need for mechanical ventilation, or death”. All the prognosis scores were also evaluated on two other outcomes that consist of “death or ICU admission” and “death”.

We evaluated *AI-severity* with several statistical measures of performance. The discriminative ability of AI-severity was of AUC = 0.78 (0.69,0.86) for the 150 left-over KB patients, and of AUC = 0.79 (0.70,0.87) for the validation IGR dataset. To compute additional measures of performance, individuals in the top tercile were assigned in a high risk group. We found that the survival function of the individuals at high risk was significantly different from the survival function of the other individuals (Figure 1, P = 4.77e-07 at KB, P = 4.00e-12 at IGR for a log-rank test). When considering a binary classification consisting of a high-risk group and a medium or low risk group, we obtained for the ‘O_2_≥15L/min or Ventilation or Death’ outcome, a positive predictive values (or precision) of 54% (0.40-0.67) (KB) and 76% (0.56-0.92) (IGR), negative predictive values of 86% (0.78-0.93) (KB) and 81% (0.73-0.88) (IGR), specificities of 75% (0.66-0.84) (KB) and 94% (0.89-0.98) (IGR), and sensitivities of 70% (0.56-0.83) (KB) and 47% (0.30-0.63) (IGR) (Table 2).

**Table 2:** Statistical measures of the performance of *AI-severity*. To compute sensitivity, specificity, PPV, and NPV, we assigned individuals into a high risk group or a low and medium risk group depending on their *AI-severity* score.The threshold to assign individuals into a high risk group was the ⅔-quantile of the *AI-severity* scores computed for patients of the KB training set. To compute measures of performance for the KB development cohort, we considered the 150 leftover individuals that were not used to learn the coefficients of *AI-severity*.

|  | AUC | Sensitivity | Specificity | PPV | NPV |
| --- | --- | --- | --- | --- | --- |
| **$O_2 \geq 15L/min$ or Ventilation or Death** |  |  |  |  |  |
| Development cohort (KB) | 0.77 (0.69-0.86) | 0.70 (0.56-0.83) | 0.75 (0.66-0.84) | 0.54 (0.40-0.67) | 0.86 (0.78-0.93) |
| Validation cohort (IGR) | 0.79 (0.70-0.87) | 0.47 (0.30-0.63) | 0.94 (0.89-0.98) | 0.76 (0.56-0.92) | 0.81 (0.73-0.88) |
| **Death or ICU** |  |  |  |  |  |
| Development cohort (KB) | 0.79 (0.71-0.86) | 0.68 (0.54-0.80) | 0.77 (0.68-0.85) | 0.60 (0.46-0.73) | 0.83 (0.74-0.90) |
| Validation cohort (IGR) | 0.86 (0.78-0.92) | 0.50 (0.34-0.65) | 0.96 (0.91-0.99) | 0.84 (0.68-0.96) | 0.81 (0.72-0.88) |
| **Death** |  |  |  |  |  |
| Development cohort (KB) | 0.81 (0.73-0.89) | 0.77 (0.60-0.91) | 0.72 (0.62-0.80) | 0.40 (0.27-0.53) | 0.92 (0.86-0.98) |
| Validation cohort (IGR) | 0.88 (0.81-0.94) | 0.62 (0.42-0.80) | 0.91 (0.84-0.96) | 0.60 (0.39-0.79) | 0.92 (0.86-0.96) |

**Figure 1:**
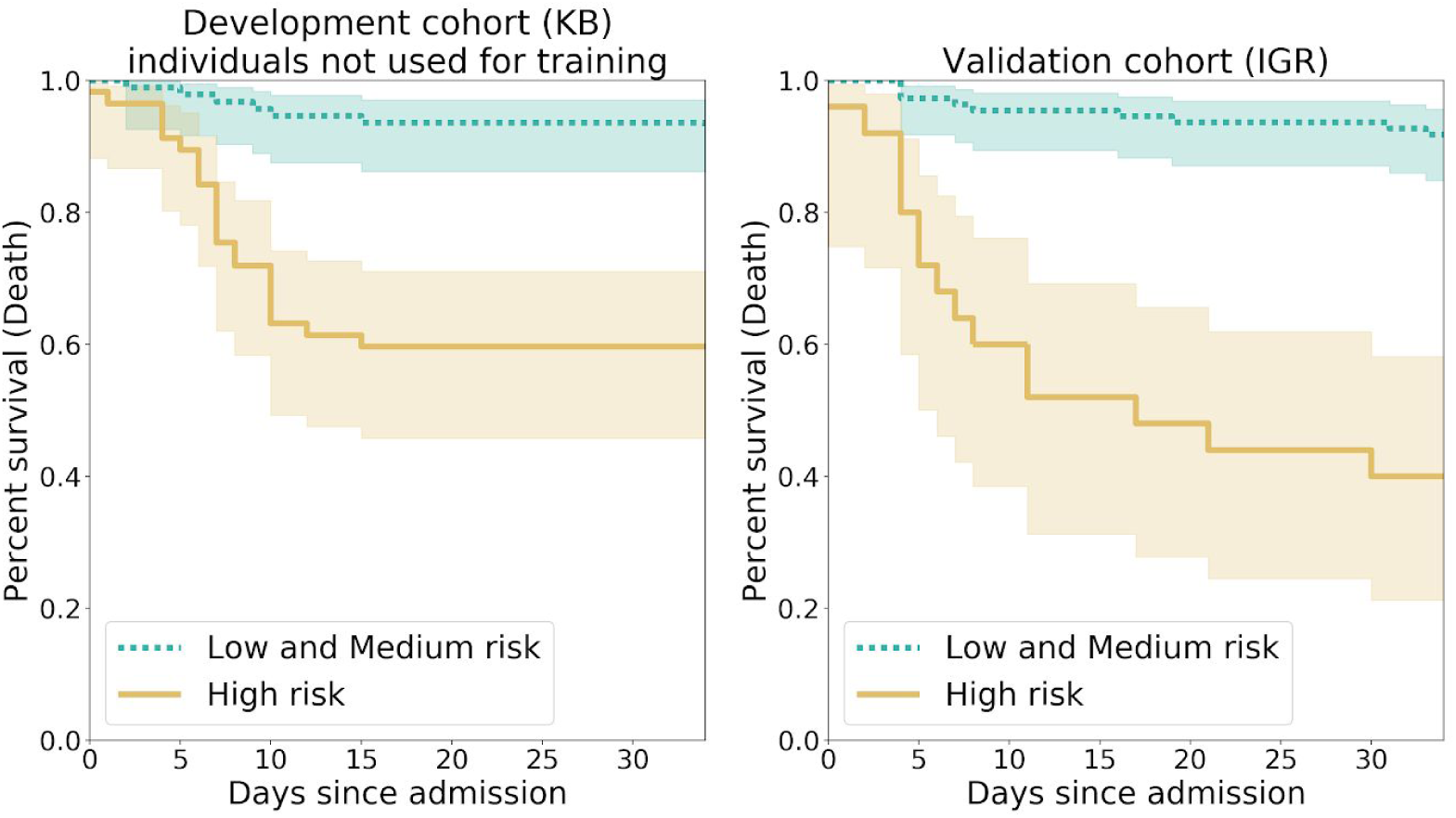
Kaplan-Meier curves for the high risk individuals and the ones with low or medium risk according to *AI-severity*. The threshold to assign individuals into a high risk group was the ⅔-quantile of the *AI-severity* score computed for patients of the KB development cohort. Kaplan-Meier curves were obtained for the 150 leftover KB patients from the development cohort (left panel) and the 135 patients of the IGR validation cohort (right panel). P-values for the log-rank `test were equal to 4.77e-07 (KB) and 4.00e-12 (IGR). The two terciles used to determine threshold values for low, medium and high risk groups were equal to 0.187, and 0.375.

*AI-severity* outperformed 11 previously published severity or mortality scores which were developed using 200 to 50,000 patients in the development and validation cohorts (Figure 2, Supp Table 3). The mean difference (averaged over outcomes) between the AUC of *AI-severity* and of other scores ranged between 0.05 (4C mortality, COVID-GRAM, CURB65, MIT analytics) and 0.16 (NEWS2) for the 150 left-over patients of the KB development cohort and between 0.07 (NEWS2 for COVID-19) and 0.28^16^ for the 135 patients of the IGR validation cohort. Among alternative scores, the COVID-GRAM, the NEWS2 for COVID-19 score, and the 4C mortality scores were the ones with the largest mean AUC values (averaged over outcomes and hospitals). The *AI-severity* score was significantly larger than the NEWS2 for COVID-19 score for all outcomes when evaluated with the left-over patients of the KB development cohort and for the ‘Death or ICU’ and the ‘Death’ outcomes when evaluated with patients from the IGR validation cohort. Differences between *AI-severity* on the one hand and the COVID-GRAM score or the 4C mortality score on the other hand were significant only for the ‘Death or ICU’ outcome when being evaluated on the left-over patients of the KB development cohort but they were significant for all outcomes when being evaluated on the validation IGR cohort.

**Figure 2:**
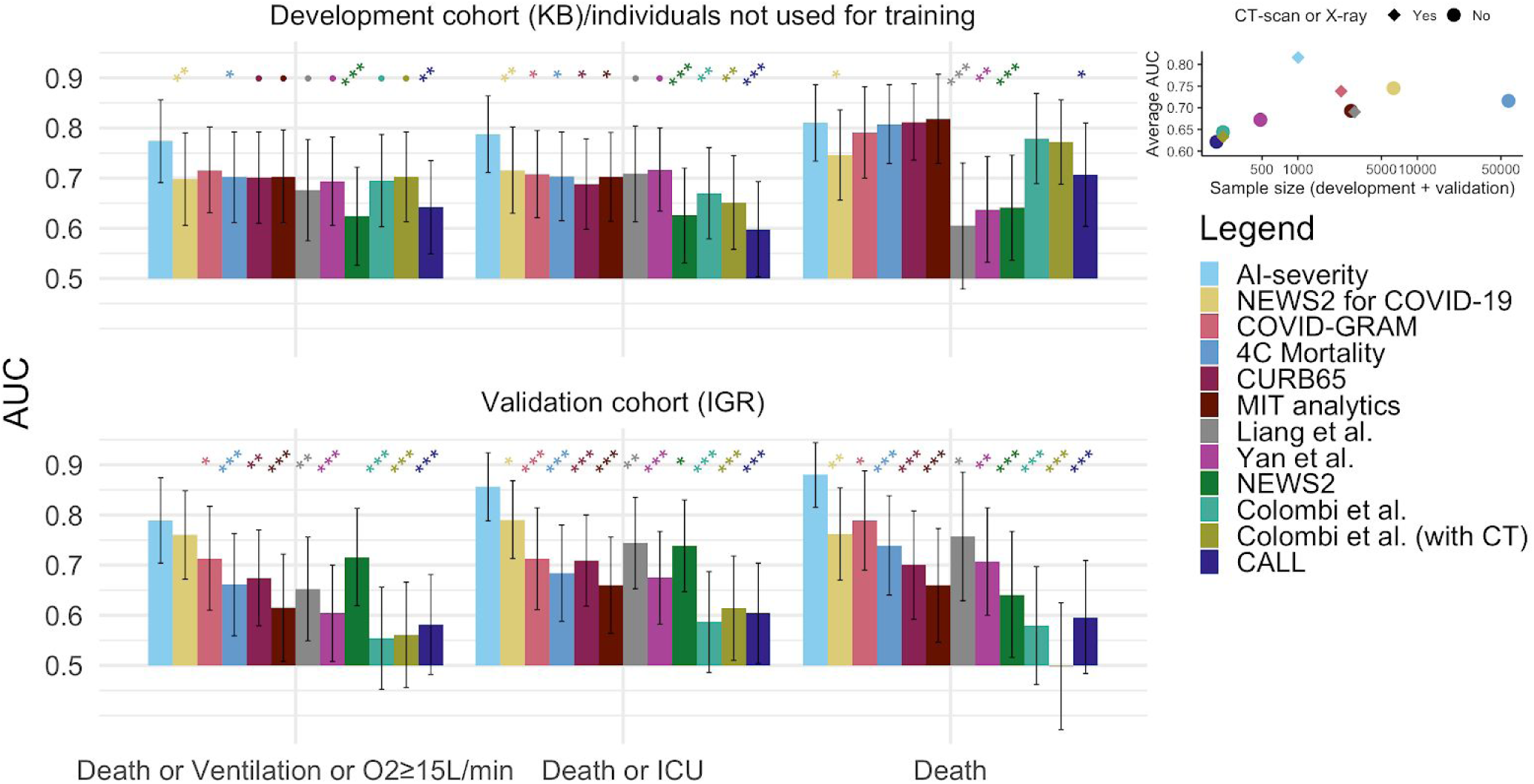
AUC values when comparing *AI-severity* to other prognostic scores for COVID-19 severity/mortality. The *AI-severity* model was trained using the severity outcome defined as an oxygen flow rate of 15 L/min or higher, the need for mechanical ventilation, or death. When evaluating *AI-severity* on the alternative outcomes, the model was not trained again. AUC results are reported on the leftover KB patients from the development cohort (150 patients) and the external validation set from IGR (135 patients). Models are sorted from left to right (and from top to bottom in the legend) by decreasing order of AUC values (averaged over outcomes and over hospitals). Error bars represent the 95% confidence intervals. The upper right inset shows the mean AUC (averaged over outcomes and over hospitals) as a function of the sample size used to construct the score, which consists of the sum of sample sizes for the development and validation cohorts. Stars indicate the order of magnitude of p-values for the DeLong procedure in which we test if AUC_*AI-severity*_ > *AUC*_*other score*_, *• 0.05<p≤0.10, * 0.01<p≤0.05, ** 0.001<p≤0.01, *** p≤0.001*.

### Development of alternative models that include CT-scan information

In addition to *AI-severity*, we considered two alternative scores that also integrate CT-scan information. The two scores include the same clinical and biological variables (age, sex, oxygen saturation, urea, platelets) than *AI-severity*. The first score (*AI-segment)* uses an automatic quantification of disease extent to include CT-scan information and the second score (*C & B & RR)* considers a radiologist quantification--available in the radiological report-instead. *AI-segment* relies on segmentation of lesions that was performed by training another deep learning model using fully annotated and partially annotated CT-scans (see Supplementary Material about segmentation). The correlation between automatic quantification of lung lesions with *AI-segment* and radiologist quantification was of 0.56 (Supp Fig 3, see Supplementary Material about segmentation).

*AI-severity* has a superior discriminative ability when compared to the alternative *C & B & RR* and *AI-segment* scores although differences of AUC were generally not significant (Supp Fig 4). The mean difference averaged over outcomes between AUC_*AI-severity*_ *and* AUC_*C & B & RR*_ (resp. AUC_*AI-segment*_) is null (resp. 0.03) for the 150 leftover KB patients of the development cohort and of 0.04 (resp. 0.01) for the IGR validation cohort. Differences between scores were not significant except when comparing *AI-severity* to *AI-segment* at KB for the ‘Death or ICU’ outcome (Supp Fig 4).

### Additional value of CT-scan information

Last, we evaluate to what extent CT-scan adds prognosis information to the clinical characteristics and biological variables from lab tests. To this end, we trained a score named *C & B* based on Clinical and Biological variables only. The AUC of the scores that integrate CT-scan information was larger or equivalent to the AUC of the *C & B* score (Supp Fig 5). The mean difference averaged over outcomes between AUC_*AI-severity*_ *and* AUC_*C&B*_ was equal to 0.03 for both cohorts. Differences between *AI-severity* and *C & B* were significant for some outcomes and cohorts but not for all combinations (Supp Fig 5). This comparative analysis shows that CT-scan adds significant prognosis information although the addition of CT-scan information increases AUC by a measurable but limited amount in both cohorts; there was a difference of AUC of 0.03 when comparing *AI-severity* to the *C & B* score.

To further investigate the added prognosis value of CT-scan, we studied the association between COVID-19 severity and the prognosis variable provided by the neural network. In the KB dataset, the three variables that were the most correlated with the prognosis variable of the neural network were oxygen saturation (r = −0.52 (−0.58,-0.48)), LDH (r = 0.46 (0.39,0.52)), CRP (r = 0.43 (0.37,0.49)) (Supp Table 4). To account for the confounding effect of these variables, we regressed the severity outcome with the neural network prognosis variable and the three correlated variables. We found that the neural network variable was significantly correlated with the severity outcome (P = 0.01). The statistical evidence for association between the neural network prognosis variable and COVID-19 severity was also found (P = 3.24 × 10^−6^) when accounting for the five additional variables of *AI-severity*. This confirms that CT-scan information captured by the neural network brings unique prognostic information.

## Discussion

Using a deep learning model to capture CT scan prognosis information, we have built the *AI-severity* score to prognose severe evolution for COVID-19 hospitalized patients. In addition to the deep learning variable, *AI-severity* is based on age, sex, oxygen saturation, urea and platelet counts. On the IGR validation cohort containing a majority of cancer patients, *AI-severity* provided values of AUCs significantly larger than the ones obtained with the best prognosis scores of our comparative analysis, which consist of COVID-GRAM, the NEWS2 score modified for COVID-19 patients and the 4C mortality score^6,12,24^. Taken together, these results show that future disease severity markers are present within routine CT scans performed at admission.

Looking back on the prognostic clinical and biological variables, we found 12 of these significantly associated with severe evolution, which is consistent with previous studies^15,25,26^. First, looking at clinical characteristics, we confirmed that male and older persons are more at risk^27^. Second, looking at clinical examination variables, we found that respiratory rate, diastolic pressure, and oxygen saturation are clinical variables associated with severity. These associations may reflect physician decisions taken for ICU triage. Inclusion criteria for critical care triage include (i) requirement for invasive ventilatory support characterized by an oxygen saturation lower than 90%, or by respiratory failure, or (ii) requirement for vasopressors characterized by hypotension and low blood pressure^28^. Third, looking at comorbidities, we confirmed the results of several meta-analyses^26,29–31^ that showed that chronic kidney disease and hypertension are linked to severity. We however did not find significant associations for other comorbidities previously associated with severity, such as diabetes, and cardiovascular diseases^31,32^. While we expected cancer patients to have more severe outcomes because they are generally older, with multiple co-morbidities and often in a treatment-induced immunosuppressive state^33–35^, we did not find this association. Several factors can explain this. Each cohort was not optimally balanced to conclusively study the association between cancer and severity: IGR admitted mostly cancer patients (80% of the patients) while KB admitted very few cancer patients (7%). Fourth, looking at COVID-19 symptoms, we did not find any significantly associated with severity. Dyspnea is a prominent symptom that has been repeatedly associated with severity and our results are compatible with a positive association with severity but we may lack a large-enough sample size to be significant^6,36,37^. Last, looking at biological measures, we found that inflammatory biomarkers, LDH and CRP are related to severity^14,25,38^. We also found association of severity with leukocytes, neutrophils and urea, the later being explained by the fact that high urea is indicative of kidney dysfunction. Thrombocytopenia (low platelet count) was not significantly associated to severity, possibly because of lack of statistical power and stringent correction for multiple testing, but association betwen thrombocytopenia and severity was in the expected direction and platelet counts are included in the 6-variable *AI-severity* score^39^.

Beyond these clinical and biological variables, chest CT-scans provided additional markers of disease severity. Significant features include the total extent of lesions, and the presence of crazy paving pattern lesions. Although the extent of disease severity and consolidation are known to be associated with severity^16,19,40–45^, our study discovered its association with crazy paving, a precursor of consolidation lesions. Initial damages to the alveoli, as well as protein and fibrous exudation, explain the early onset of GGO. As the disease progresses, more and more inflammatory cells infiltrate the alveoli and interstitial space, followed by diffuse alveolar lesions and the formation of a hyaline membrane, which results in a crazy paving appearance, which is then followed by consolidation on the CT examination^46,47^.

Compared to a radiologist’s reporting and quantification of lesions, there are several advantages to capturing CT-scan information through a deep learning model. Good reproducibility is a key element for imaging biomarkers, and visual inspection of images introduces variability that can hinder its clinical application^48^. Another advantage is that radiologists are faced with the challenge that large numbers of cases must be read, annotated and prioritized in a COVID-19 pandemic. AI analysis of radiological images has the potential to reduce this burden and speed up their reading time. Finally, prognosis scores obtained with deep learning models trained on CT-scans are more predictive of severity than a quantification of disease extent performed by a radiologist. We indeed showed that internal representation of the *AI-severity* neural network model captures clinical information from CT scans, and this can be particularly useful when some clinical or lab measurements are missing.

Our reported prognostic values for CT-scan-based models (AUC range of 0.70 - 0.80) are lower than the 0.85 AUC reported in a previously published study that uses deep learning with CT-scan images for prognosis^17^. We hypothesize that this is due to use of different outcome definitions, as well as different patient characteristics in the study cohorts (age, severity at admission, etc). Hospital admission criteria vary between countries and hospitals; for instance, the proportion of deaths in our French KB and IGR cohorts was of 16-17%, while it was of 39% in the study that reported larger AUC values^17^. When applying other previously published scores to the KB and IGR datasets, we found smaller AUC scores than reported values in the original papers. This difference can again be explained by differing patient characteristics, and different measures of severity between studies^7,10,166,9^

Our evaluation of *AI-severity* and of alternative scores revealed that including CT scan information in addition to clinical and biological information significantly improves prognosis of future severity at least for the IGR validation cohort. However, the neural network prognosis variable was correlated to biological and clinical severity biomarkers such as CRP levels, tissue damage (LDH) and oxygenation—highlighting some information redundancy between data modalities—explaining the relatively modest gain of AUC provided by CT-scan36,49–51.

Beyond AI modeling, our study shows that the 6-variable *AI-severity* score integrating a radiological quantification of lesions with key clinical and biological variables provides accurate severity predictions, and can rapidly become a reference scoring approach for hospitalized patients.

## Methods

### Description of the retrospective study

Data including CT-scans, were collected at two French hospitals (Kremlin Bicêtre Hospital, APHP, Paris denoted as KB and Gustave Roussy Hospital, Villejuif denoted as IGR). CT scans, clinical, and biological data were collected in the first 2 days after hospital admission. This study has received approval of ethic committees from the two hospitals and authors submitted a declaration to the National Commission of Data Processing and Liberties (N° INDS MR5413020420, CNIL) in order to get registered in the medical studies database and respect the General Regulation on Data Protection (RGPD) requirements. An information letter was sent to all patients included in the study. We stopped to update information about patient status on the 5th of May. Among the 1,003 patients of the study, two patients asked to be excluded from the study.

Inclusion criteria were (1) date of admission at hospital (from the 12th of February to the 20th of March at Kremin Bicêtre and from the 2nd of March to the 24th of April at Institut Gustave Roussy) and (2) a positive diagnosis of COVID-19. Patients were considered positive either because of a positive RT-PCR (real-time fluorescence polymerase chain reaction) based on nasal or lower respiratory tract specimens or a CT scan with a typical appearance of COVID-19 as defined by the ACR criteria for negative RT-PCR patients^52^. Children and pregnant women were excluded from the study.

The clinical and laboratory data were obtained from detailed medical records, cleaned and formatted retrospectively by 10 radiologists with 3 to 20 years of experience (5 radiologists at GR and 5 at KB). Data include demographic variables: age and sex, variables from the clinical examination include: body weight and height, body mass index, heart rate, body temperature, oxygen saturation, blood pressure, respiratory rate, and a list of symptoms including cough, sputum, chest pain, muscle pain, abdominal pain or diarrhoea, and dyspnea. Health and medical history data include presence or absence of comorbidities (systemic hypertension, diabetes mellitus, asthma, heart disease, emphysema, immunodeficiency) and smoker status. Laboratory data include conjugated alanine, bilirubin, total bilirubin, creatine kinase, CRP, ferritin, haemoglobin, LDH, leucocytes, lymphocyte, monocyte, platelet, polynuclear neutrophil, and urea.

### CT scan acquisition

CT scan data were available for 980 patients representing a total of 506,341 2D images (517 slices per patient on average). Summary statistics for the clinical, biological, and CT scan data are provided in Table 1. Three different models of CT scanners were used : two General Electric CT scanners (Discovery CT750 HD and Optima 660 GE Medical Systems, Milwaukee, USA) and a Siemens CT scanner (Somatom Drive; Siemens Medical Solutions, Forchheim). All patients were scanned in a supine position during breath-holding at full inspiration. The acquisition and reconstruction parameters were of 120kV tube voltage with automatic tube current modulation (100-350 mAs), 1 mm slice thickness without interslice gap, using filtered-back-projection (FBP) reconstruction (SOMATOM Drive) or blended FBP/iterative reconstruction (Discovery or Optima). Axial images with slice thickness of 1 mm were used for coronal and sagittal reconstructions.

### Radiology reports

COVID-19 associated CT imaging features were obtained from radiologist reports that follow the guidelines of several scientific societies of radiology (French SFR, STR, ACR, RSNA) regarding the reporting of chest CT findings related to COVID-19 ^52^. The template of the radiologist report (https://ebulletin.radiologie.fr/actualites-covid-19/compte-rendu-tdm-thoracique-iv-0) was available the 17th of March and the reports were completed retrospectively for the patients who were admitted to the hospital before that date. CT imaging characteristics were evaluated to provide the five following variables : (i) ground glass opacity (GGO) (rounded / non rounded / absent) that is defined as an increase in lung density not sufficient to obscure vessels or preservation of bronchial and vascular margins, (ii) consolidation (rounded / non rounded / absent) that occurs when parenchymal opacification is dense enough to obscure the vessels’ margins and airway walls and other parenchymal structures, (iii) the crazy-paving pattern (present/absent) that is defined as ground-glass opacification with associated interlobular septal thickening^53^, (iv) peripheral topography (present/absent) that corresponds to the spatial distribution of lesions in the one-third external part of the lung, and (v) inferior predominance (present/absent) that is defined as a predominance of lesions located in the lower segments of the lung. A rounded pattern (for GGO and consolidation) is defined as a lesion presenting a well delineated shape. In addition to the five CT imaging features, radiologists assessed the extent of lung lesions according to the evaluation criteria established by the French Society of Radiology (SFR)^54^. Disease extent can be: absent / minimal (<10%) / moderate (10-25%) / extensive (25-50%) / severe (>50%) / critical >75%. The coding absent / minimal / moderate extensive / severe / critical was based on a quantitative variable with values of 0 / 1 / 2 / 3 / 4 / 5. Variables were automatically extracted from the report using optical character recognition.

### Statistical analysis

When detecting association with the severity outcome, odds ratio and P-values (two-sided tests) were computed separately for each hospital using logistic regression (*glm* function of the R statistical software). P-values from the two different hospitals were pooled using the Stouffer meta-analysis formula accounting for the two different sample sizes. For association between severity and each variable, we considered Bonferroni correction accounting for 58 variables. To compute confidence intervals for AUC values, we considered DeLong method^55^. Survival functions were obtained using Kaplan-Meier estimators.

### Deep learning models for severity classification based of CT scans

The deep learning model was defined as an ensemble of two sub-models, as illustrated in Supp Fig 2. Each sub-model predicted disease severity from CT scans without using any expert annotations at the slice level. Preprocessing of the data consisted of resizing the CT scans to a fixed pixel spacing of (0.7mm, 0.7mm, 10mm) and applying a specific windowing on the HU intensities. Each sub-model is composed of two blocks: a deep neural network called *feature extractor* and a penalized logistic regression. The two sub-models feature extractors are EfficientNet-B0^56^ pre-trained on the ImageNet public database and ResNet50^57^ pre-trained with MoCo v2^58^ on one million CT scan slices from both Deep Lesion^59^ and LIDC^60^. Each of these networks provide an embedding of the slices of the input CT scans into a lower-dimensional feature space (1280 for EfficientNet-B0 and 2048 for ResNet50). For the ResNet50-based sub-model, we reduced the dimension of the feature space using a principal component analysis with 40 components before applying logistic regression. A different windowing was applied on the CT scans before the feature extractor : (−1000 HU, 600 HU) for EfficientNet-B0 and (−1000 HU, 0 HU), (0 HU, 1000 HU) and (−1000 HU, 4000 HU) for ResNet50. Predictions of *AI-severity* were obtained by averaging predictions of the submodels using equal weights. Optimisation of the architecture of the network (preprocessing, feature extraction or model architecture and training, feature engineering, model aggregation) was performed using a 5 fold cross validation on the training set of 646 patients from KB.

CT scans may contain devices such as catheters (EKG monitoring, oxygenation tubing…) that are easily detectable in a CT and can bias prediction of severity. Indeed, there is a risk of detecting the presence of a technical device associated with severity instead of detecting the radiological features associated with severity^61^. In order to ensure that medical devices do not affect feature extraction, all voxels outside of the lungs were masked using a pre-trained U-Net lung segmentation algorithm^62^.

### Multivariate models to predict severity

The different models that combine multiple features to predict severity were fitted using logistic regression (*AI-severity, AI-segment, C & B, C & B & RR*). Models were trained using cross validation with 5 folds on the training dataset of 646 patients from KB, and folds were stratified by age and severity outcome. Variables that were available for less than 300 patients of the training set (conjugated bilirubin and alanine) were not used. For the remaining variables, missing values were imputed by the average over patients of the training set. L2 regularization was applied when fitting logistic regression. The regularization coefficient was determined by maximizing the average AUC over the 5 cross-validation folds, using a range of different values ranging from 0.01 to 100. XGBoost algorithm was also evaluated but did not show better performance than logistic regression. We use pandas and scikit-learn to manipulate data, train and evaluate machine learning algorithms^63^.

To select variables in the multivariate models, we considered a forward feature selection technique (Supp Fig 6). The first variable included in the model is the variable which provides the largest AUC values. Then, we computed AUC values for all models with two variables including the first one that has already been included. We continued this procedure until all variables were included. Performances of the models increased quickly when the first variables were included and then AUC values reached a plateau (Supp Fig 6). We used the elbow method to select the parsimonious set of variables that is found when a plateau of AUC is reached. For the three models that include CT-scan information, we consider the model *C & B & RR* to perform variable selection. The three models (*AI-severity, C & B & RR, AI-segment*) were then trained using the six variables found with our variable selection procedure. The variable selection procedure for the *C & B* model that contains clinical and biological variables only indicated that 10 variables should be kept in the model (see Supp Table 5 for the list of selected variables).

### Other scores to predict severity and mortality

We performed a comparison of the proposed AI-severity model with 11 other COVID-19 severity scores published in the literature: COVID-Gram^6^, 2 scores from Colombi et al.^16^, CALL^9^, CURB-65^64^, Yan et al.^7^, Liang et al.^65^, NEWS2 and NEWS2 for COVID-19^66^, 4C mortality score^12^, and MIT analytics (https://www.covidanalytics.io/mortality_calculator). Among these scores, only three included radiological information in their model: the presence of an X-ray abnormality for COVID-Gram and Liang et al., and the lung disease extent for Colombi et al.. The number of considered clinical and biological variables for these scores varied (from 3 for Yan et al. to 14 for NEWS2 for COVID), as well as the model architecture (simple scoring system, logistic regression, xgboost or multilayer perceptron), and the outcome they were trained on (such as death or admission to ICU). Notable variation between scores include the definition of comorbidities and details about how it has been computed for the different scores are provided in Supp Table 6. For missing variables, we manually imputed the missing variables with a constant value (Supp Table 6). Due to the poor performances of one of the score^7^, we retrained their score by repeating their training procedure with the KB development cohort.

## Supporting information

Main supplementary material

Supplementary material about segmentation

## Data Availability

The dataset of patients hospitalized at Kremlin-Bicetre (KB) and Institut Gustave Roussy (IGR) are stored on a server at Institut Gustave Roussy (IGR). The data are available from the first author upon request subject to ethical review.

## Data Availability

The dataset of patients hospitalized at Kremlin-Bicêtre (KB) and Institut Gustave Roussy (IGR) are stored on a server at Institut Gustave Roussy (IGR). The data are available from the first author upon request subject to ethical review.

## Code Availability

Code to execute *AI-severity* as well as the other models we developed and the 11 additional models we evaluated are available online at https://github.com/owkin/scancovia.

## Acknowledgements

We would like to thank J.-Y. Berthou, H. Berry, and Ph. Gesnouin from Inria and M. He, R. Patel, G. Rouzaud, B. Schmauch, J. Du Terrail from Owkin and F Lion from Gustave Roussy for their support. This work was granted COVID-19 priority access to the HPC resources of IDRIS (Jean Zay) under the allocation AD011011728 made by GENCI.

## Author Contributions

N.L., S.A., E.C.,P.H.,R.M.,N.L.,P.T., E.B.,M.S., A.S., F.C.,S.J., M.S., I.B., J.D.,JC.P., H.T.,E.P.,G.W., T.C., F.B.,MF.B.,M.B conceived the idea of this paper

N.L., S.A., E.C., H.G.,P.H., M.D., S.S., O.M., MP.T., JP.L.,R.M.,N.L.,P.T., E.B.,G.G, C.B.,S.J., F.G.,N.T.,Y.L., T.D., K.G., A.N., M.T., S.V., M.S., I.B., Y.B, E.P., M.A., J.D.,F.B., A.G.,J.D.,JC.P., H.T.,E.P.,G.W., T.C., F.B.,MF.B.,M.B participated to the acquisition and treatment of data

N.L., S.A., E.C.,P.H.,R.M.,N.L.,P.T., E.B.,S.J., M.S., P.J., I.B., J.D.,JC.P.,H.T.,E.P.,G.W., T.C., MF.B.,M.B.implemented the analysis

N.L., S.A., E.C.,P.H.,R.M.,N.L.,P.T., E.B.,S.J., M.S., I.B., J.D.,JC.P., H.T.,E.P.,G.W., T.C., MF.B.,M.B.contributed to the writing of the manuscript

## Competing Interests statement

The authors declare the following competing interests:

- Employment: Michael Blum, Paul Herent, Rémy Dubois, Nicolas Loiseau, Paul Trichelair, Etienne Bendjebbar, Simon Jégou, Meriem Sefta, Paul Jehanno, Fabien Brulport, Olivier Dehaene, Jean-Baptiste Schiratti, Kathryn Schutte, Elodie Pronier, Jocelyn Dachary, Adrian Gonzalez, employed by Owkin
- Co-founders of Owkin Inc : Thomas Clozel, Gilles Wainrib.

## Notes

### Funding Statement

Owkin employees are paid by Owkin, Inc.

### Author Declarations

This study has received approval of ethic committees from the two hospitals and authors submitted a declaration to the National Commission of Data Processing and Liberties (Number INDS MR5413020420, CNIL) in order to get registered in the medical studies database and respect the General Regulation on Data Protection (RGPD) requirements.

### Summary of Updates

Ai-severity is now the only score we developed to be put forward. Comprehensive comparison with 11 scores that predict severity/mortality.

