## Supplementary material for "Integration of clinical characteristics, lab tests and a deep learning CT scan analysis to predict severity of hospitalized COVID-19 patients": Main supplementary material

### Supplementary Figures and Tables

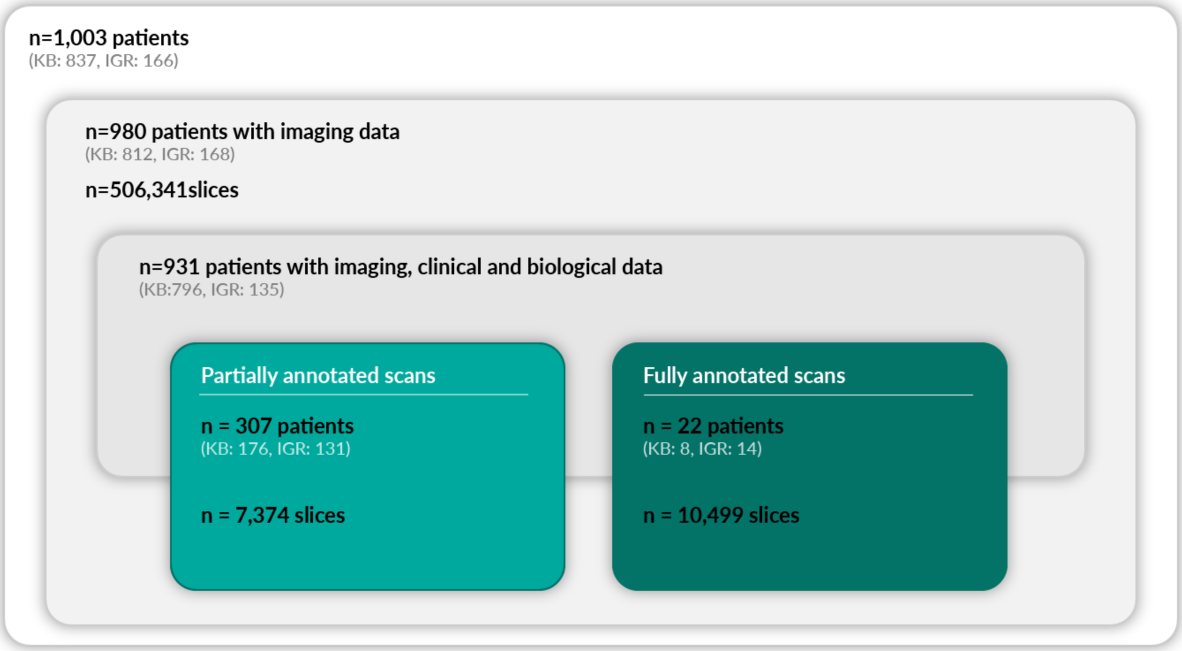


**Supp Fig 1: Description of the retrospective cohort.** Number of patients and repartition per hospital for different all patients, patients

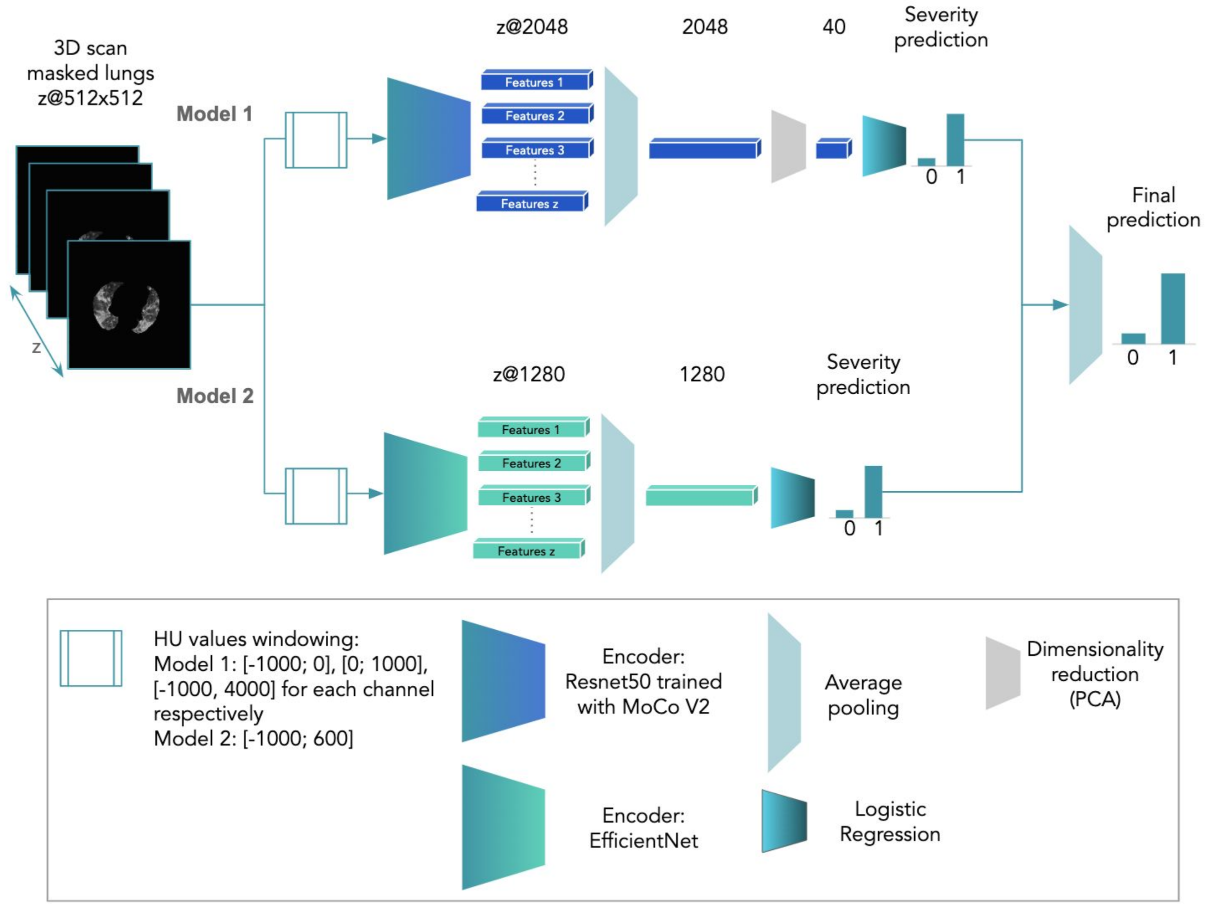


**Supp Fig 2: Neural network to predict severity from 3D chest CT scans.** The final prediction of the network is one of the 6 variables of the *AI-severity* score. Two different pipelines were used: one using Resnet50 (trained with MocoV2 on 1 million public CT scan slices) as encoder (model 1) and one using EfficientNet B0 as encoder (model 2).

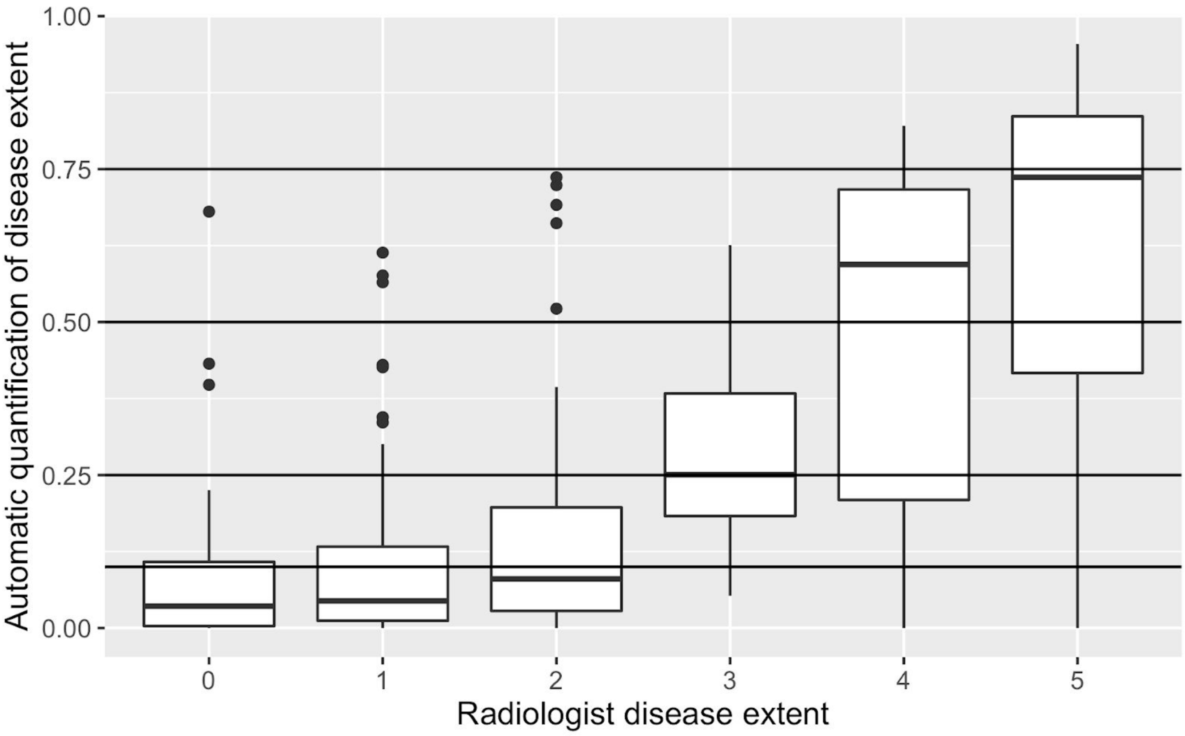


**Supp Fig 3: Boxplot to compare automatic quantification of disease extent using a neural network segmentation model and disease extent as quantified by a radiologist.** The coding of disease extent in the radiologist report is as follows: 0 (0% of lesions), 1 (<10% of lesions), 2 (between 10 and 25% of lesions), 3 (between 25 and 50% of lesions), 4 (between 50 and 75% of lesions), 5 (more than 75% of lesions). The lower and upper hinges correspond to the first and third quartiles. The upper whisker extends from the hinge to the largest value no further than 1.5 * IQR from the hinge (where IQR is the inter-quartile range). The lower whisker extends from the hinge to the smallest value at most 1.5 * IQR of the hinge. Data beyond the end of the whiskers are called "outlying" points and are plotted individually.

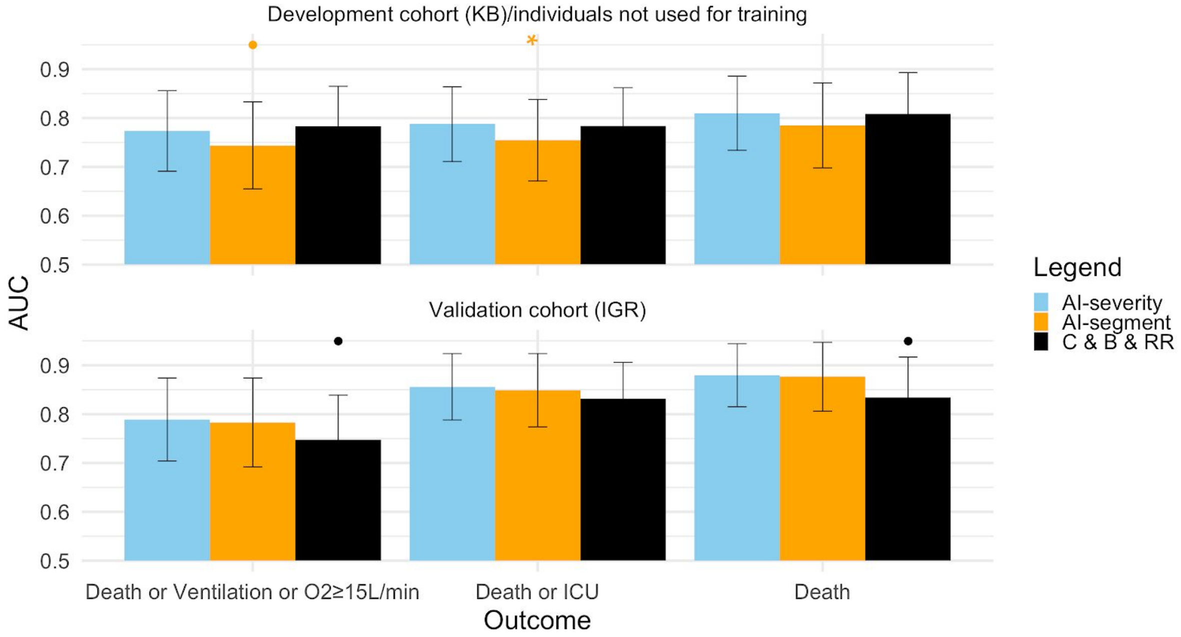


**Supp Fig 4**: **AUC values when comparing *AI-severity* to *AI-segment* and to the model including Clinical, Biological variables and disease extent extracted from a Radiologic Report *(C & B & RR)*.** All three models were trained using the severity outcome defined as an oxygen flow rate of 15 L/min or higher, the need for mechanical ventilation, or death. When evaluating the three models on the alternative outcomes, models were not trained again. AUC results are reported on the leftover KB patients from the development cohort (150 patients) and the external validation set from IGR (135 patients). Error bars represent the 95% confidence intervals. Stars indicate the order of magnitude of p-values for the DeLong procedure in which we test if $AUC_{AI\text{-}severity} > AUC_{other\ score}$, *• 0.05<p≤0.10, * 0.01<p≤0.O5, ** 0.001<p≤0.01, *** p≤0.001.*

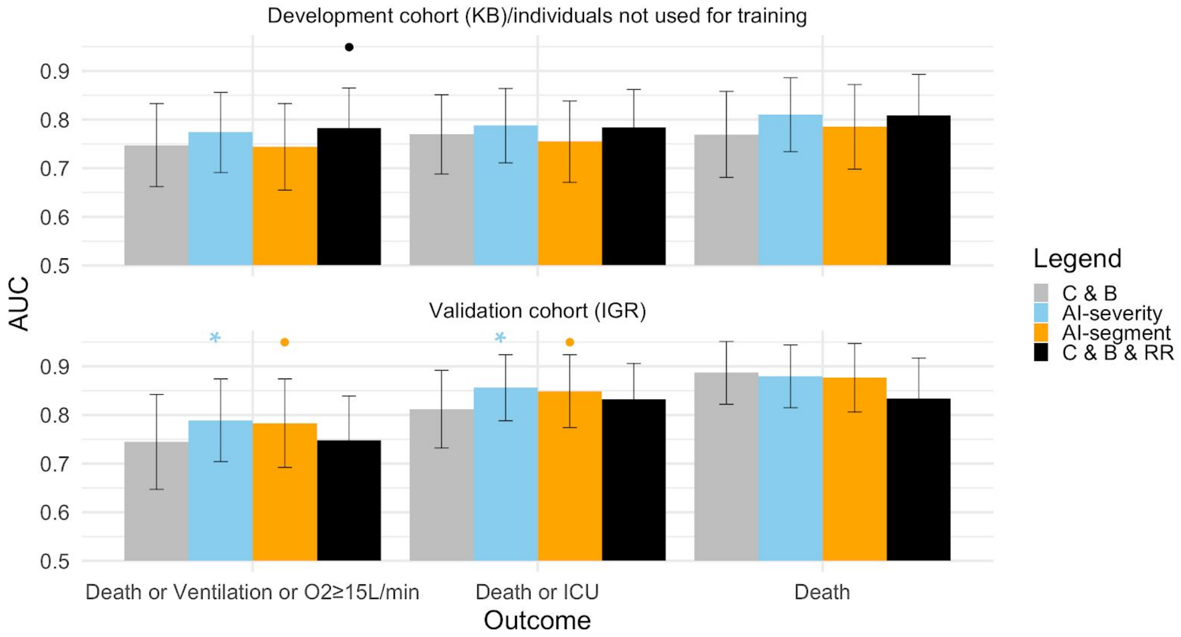


**Supp Fig 5**: **AUC values when comparing the model that includes Clinical, and Biological variables (*C & B*) to the three models that additionally include CT-scan information (*AI-severity*, *C & B & RR*, *AI-segment*).** All four models were trained using the severity outcome defined as an oxygen flow rate of 15 L/min or higher, the need for mechanical ventilation, or death. When evaluating the three models on the alternative outcomes, models were not trained again. AUC results are reported on the leftover KB patients from the development cohort (150 patients) and the external validation set from IGR (135 patients). Error bars represent the 95% confidence intervals. Stars indicate the order of magnitude of p-values for the DeLong procedure in which we test $AUC_{C\ \&\ B} < AUC_{other\ score}$, *• 0.05<p≤0.10, * 0.01<p≤0.05, ** 0.001<p≤0.01, *** p≤0.001.*

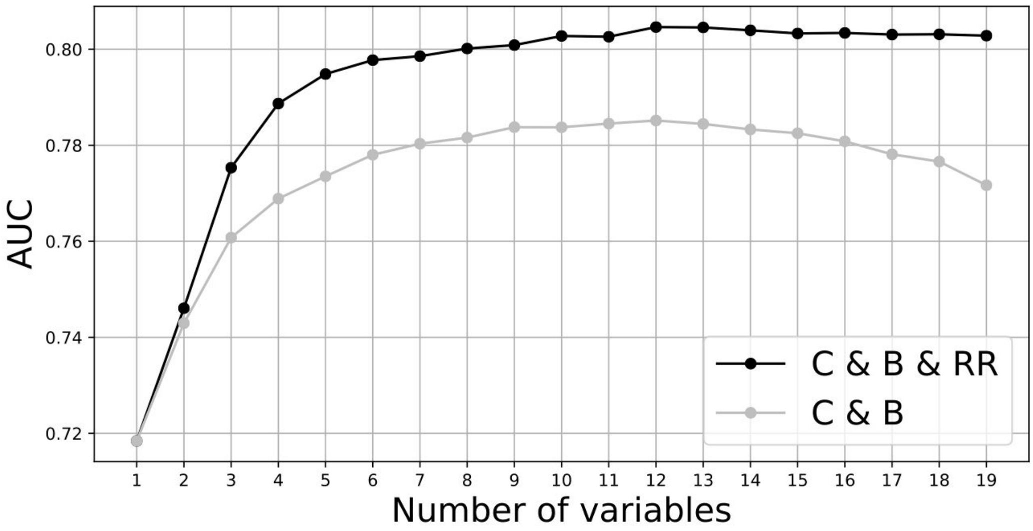


**Supp Fig. 6: AUC curve as a function of the number of clinical and biological information added to the multimodal model.** Variables included in the models consist of CT scan variables only and then a greedy algorithm adds clinical or biological variables iteratively. At each step of the algorithm, the variable that results in the largest increase of AUC score is added.

| Variable | AUC for the 150 leftover patients of the KB development cohort | AUC for the 135 IGR patients of the validation cohort |
|---|---|---|
| Age > 60 | 0.884 (0.828 - 0.940) | 0.786 (0.710 - 0.862) |
| Sex | 0.933 (0.892 - 0.975) | 0.893 (0.838 - 0.947) |
| Oxygen saturation > 90 | 0.761 (0.681 - 0.840) | 0.782 (0.676 - 0.888) |
| Disease extent > 2 | 0.926 (0.887 - 0.965) | 0.881 (0.819 -0.943) |
| Crazy paving | 0.775 (0.700 - 0.851) | 0.725 (0.637 - 0.812) |
| Condensation | 0.6365 (0.534 - 0.737) | 0.675 (0.583 -0.767) |
| GGO | 0.800 (0.655 - 0.944) | 0.583 (0.475 - 0.690) |

**Supp Table 1**: ***AI-severity* model performances on other classification tasks than severity prediction.** AUC scores are reported on both KB and IGR validation sets when re-training the *AI-severity* model to predict a few clinical and radiological variables we have selected. To retrain the model, we considered the last hidden layer of *AI-severity* as a feature vector and used logistic regression to predict clinical/radiological outcome.

| Variable | Coding/unit | Transformation | Coefficient |
|---|---|---|---|
| Oxygen saturation | % | -log(1 + 100 - X) | -0.569 |
| Neural network variable | | None | 0.769 |
| Age | year | None | 0.0121 |
| Sex | 1 for male<br>0 for female | None | 0.412 |
| Platelet | G/L | log(0.001 + X) | -0.567 |
| Urea | mmol/L | log(0.001 + X) | 0.393 |

**Supp Table 2: Coefficients, transformation, and units to compute the *AI-severity* score.**

| Model description | KB | IGR | KB CV |
|---|---|---|---|
| O2≥15L/min or Ventilation or Death | | | |
| AI-severity | 0.774 (0.691 - 0.856) | **0.789 (0.704 - 0.874)** | **0.793 (0.689 - 0.881)** |
| Neural network analysis | 0.763 (0.674 - 0.851) | 0.748 (0.657 - 0.839) | 0.748 (0.627 - 0.839) |
| C & B | 0.747 (0.662 - 0.833) | 0.745 (0.647 - 0.842) | 0.776 (0.686 - 0.871) |
| C & B & RR | **0.783 (0.702 - 0.865)** | 0.748 (0.658 - 0.839) | **0.793 (0.713 - 0.899)** |
| AI-segment | 0.744 (0.655 - 0.833) | 0.783 (0.692 - 0.874) | 0.787 (0.685 - 0.881) |
| MIT analytics | 0.703 (0.611 - 0.796) | 0.615 (0.508 - 0.722) | |
| CALL | 0.642 (0.549 - 0.735) | 0.582 (0.482 - 0.681) | |
| Colombi et al. | 0.695 (0.603 - 0.787) | 0.554 (0.452 - 0.656) | |
| Colombi et al. (with CT scan) | 0.702 (0.613 - 0.792) | 0.561 (0.456 - 0.666) | |
| COVID_GRAM | 0.716 (0.631 - 0.802) | 0.713 (0.610 - 0.817) | |
| CURB65 | 0.701 (0.610 - 0.792) | 0.674 (0.579 - 0.770) | |
| Yan et al. | 0.694 (0.606 - 0.782) | 0.604 (0.508 - 0.700) | |
| NEWS2_carr | 0.624 (0.526 - 0.722) | 0.716 (0.619 - 0.813) | |
| NEWS2 for COVID-19 | 0.698 (0.606 - 0.790) | 0.760 (0.672 - 0.848) | |
| 4C mortality | 0.702 (0.611 - 0.792) | 0.661 (0.559 - 0.763) | |
| Liang et al. | 0.676 (0.575 - 0.777) | 0.652 (0.549 - 0.756) | |
| Death or ICU | | | |
| AI-severity | **0.788 (0.711 - 0.864)** | **0.856 (0.788 - 0.924)** | 0.793 (0.708 - 0.890) |
| Neural network analysis | 0.767 (0.685 - 0.849) | 0.825 (0.748 - 0.902) | 0.749 (0.632 - 0.857) |
| C & B | 0.770 (0.688 - 0.851) | 0.812 (0.732 - 0.892) | 0.782 (0.679 - 0.878) |
| C & B & RR | 0.784 (0.706 - 0.862) | 0.832 (0.757 - 0.906) | **0.794 (0.714 - 0.892)** |
| AI-segment | 0.755 (0.671 - 0.838) | 0.849 (0.774 - 0.924) | 0.792 (0.715 - 0.900) |
| MIT analytics | 0.703 (0.614 - 0.791) | 0.660 (0.564 - 0.756) | |
| CALL | 0.598 (0.503 - 0.693) | 0.604 (0.504 - 0.704) | |
| Colombi et al. | 0.670 (0.579 - 0.761) | 0.587 (0.486 - 0.687) | |
| Colombi et al. (with CT scan) | 0.651 (0.558 - 0.745) | 0.614 (0.510 - 0.718) | |
| COVID_GRAM | 0.708 (0.621 - 0.795) | 0.713 (0.611 - 0.814) | |
| CURB65 | 0.688 (0.598 - 0.778) | 0.709 (0.618 - 0.800) | |
| Yan et al. | 0.717 (0.634 - 0.800) | 0.675 (0.582 - 0.767) | |
| NEWS2_carr | 0.626 (0.531 - 0.720) | 0.739 (0.647 - 0.830) | |
| NEWS2 for COVID-19 | 0.716 (0.630 - 0.802) | 0.790 (0.713 - 0.868) | |
| 4C mortality | 0.704 (0.615 - 0.792) | 0.684 (0.588 - 0.780) | |
| Liang et al. | 0.709 (0.613 - 0.804) | 0.744 (0.653 - 0.835) | |
| Death | | | |
| AI-severity | 0.810 (0.734 - 0.886) | **0.880 (0.815 - 0.944)** | 0.787 (0.718 - 0.902) |
| Neural network analysis | 0.752 (0.660 - 0.845) | 0.753 (0.644 - 0.862) | 0.710 (0.630 - 0.835) |
| C & B | 0.769 (0.681 - 0.858) | 0.887 (0.822 - 0.951) | **0.794 (0.683 - 0.914)** |
| C & B & RR | 0.809 (0.726 - 0.893) | 0.834 (0.752 - 0.917) | 0.786 (0.685 - 0.924) |
| AI-segment | 0.785 (0.698 - 0.872) | 0.877 (0.806 - 0.947) | 0.782 (0.711 - 0.916) |
| MIT analytics | **0.818 (0.729 - 0.907)** | 0.660 (0.546 - 0.773) | |
| CALL | 0.707 (0.604 - 0.810) | 0.596 (0.484 - 0.709) | |
| Colombi et al. | 0.779 (0.689 - 0.869) | 0.579 (0.462 - 0.697) | |
| Colombi et al. (with CT scan) | 0.772 (0.688 - 0.856) | 0.498 (0.372 - 0.625) | |
| COVID_GRAM | 0.791 (0.700 - 0.882) | 0.789 (0.690 - 0.888) | |
| CURB65 | 0.812 (0.736 - 0.888) | 0.700 (0.592 - 0.808) | |
| Yan et al. | 0.637 (0.532 - 0.743) | 0.707 (0.600 - 0.814) | |
| NEWS2_carr | 0.641 (0.536 - 0.746) | 0.641 (0.516 - 0.767) | |
| NEWS2 for COVID-19 | 0.746 (0.656 - 0.836) | 0.762 (0.670 - 0.854) | |
| 4C mortality | 0.807 (0.729 - 0.886) | 0.739 (0.640 - 0.838) | |
| Liang et al. | 0.605 (0.479 - 0.730) | 0.757 (0.629 - 0.885) | |

**Supp Table 3**: **AUC values for the different models on the different sets.** Each model (Neural network analysis, *AI-severity*, *C & B*, *C & B & RR*, *AI-segment*) was trained on 646 patients from KB. Results are reported on the leftover 150 patients from the development KB cohort and for the 135 patients from the IGR validation set. For the models we trained, we also report performance obtained using 5 fold cross validation stratified by outcome and age (CV KB). For each outcome, the best models are highlighted in boldface. C & B: Clinical and Biological variables, C & B & RR: Clinical and Biological variables and the ones of the Radiological Report.

| | Correlation | Lower 95% C.I. | Upper 95% C.I. |
|---|---|---|---|
| Disease extent | 0.62 | 0.58 | 0.67 |
| Oxygen saturation | -0.53 | -0.58 | -0.48 |
| LDH | 0.46 | 0.39 | 0.52 |
| CRP | 0.43 | 0.37 | 0.49 |
| Age | 0.3 | 0.24 | 0.36 |
| Respiratory rate | 0.25 | 0.17 | 0.32 |
| Neutrophil | 0.25 | 0.18 | 0.31 |
| Leucocytes | 0.22 | 0.15 | 0.29 |
| Urea | 0.18 | 0.11 | 0.25 |
| Ferritin | 0.15 | 0.03 | 0.26 |
| Diastolic pressure | -0.15 | -0.22 | -0.07 |
| BMI | 0.14 | 0.04 | 0.23 |
| Total bilirubin | 0.11 | 0.03 | 0.18 |
| Platelet | 0.09 | 0.02 | 0.16 |
| Weight | 0.08 | -0.01 | 0.16 |
| Creatine kinase | 0.07 | -0.01 | 0.15 |
| Haemoglobin | -0.06 | -0.14 | 0.01 |
| Body temperature | 0.06 | -0.01 | 0.13 |
| Cardiac frequency | -0.06 | -0.13 | 0.02 |
| Conjugated bilirubin | -0.05 | -0.32 | 0.23 |
| Lymphocyte | -0.04 | -0.11 | 0.03 |
| Systolic pressure | -0.04 | -0.11 | 0.03 |
| Symptoms duration before examination | -0.04 | -0.11 | 0.03 |
| Monocyte | -0.02 | -0.09 | 0.05 |
| Height | -0.01 | -0.11 | 0.09 |

**Supp Table 4**: **Correlation of clinical and biological variables with the prognosis obtained with a weakly-supervised neural network.** Correlation was computed using 817 patients from the KB hospital. Variables are sorted in decreasing order when considering the squared correlation value for ranking.

| **Models** | **Variables included** | | | | | | | | | |
|---|---|---|---|---|---|---|---|---|---|---|
| *1. AI-severity*<br>*2. AI-segment*<br>3. Clinical & bio & Radiological Report (*C & B & RR*) | Oxygen saturation | Disease extent | Age | Sex | Platelet | Urea | | | | |
| Clinical and bio (C & B) | Oxygen saturation | Age | Sex | LDH | Platelet | Chronic kidney disease | Dyspnea | Hypertension | Neutrophil | Urea |

**Supp Table 5**: **Names of the variables included in the different models.**

| Score | Variable | Value used |
|---|---|---|
| CALL score | Comorbidity | 1 if any of cardiac disease, asthma, emphysema, diabetes, or hypertension else 0 |
| Colombi et al. | Cardiovascular disease | 1 if any of cardiac disease or hypertension else 0 |
| COVID-Gram and Liang et al. | XRay abnormality | 1 if any lesion is observed on the CT scan, else 0 |
| COVID-gram | Hemoptysis | 0 |
| COVID-gram, NEWS2 | Unconsciousness | 0 |
| COVID-gram | Number of comorbidities | Count of cardiac disease, asthma, diabetes, emphysema, chronic kidney disease, hypertension |
| Liang et al. | Number of comorbidities | Count of cardiac disease, diabetes, emphysema, chronic kidney disease, cancer, hypertension |
| Liang et al. | Chronic obstructive pulmonary disease | Same value as emphysema |
| 4C | Number of comorbidities | Count of cardiac disease, diabetes, emphysema, chronic kidney disease, cancer |
| 4C | Glasgow coma score | 0 |
| NEWS2 | Air or oxygen | 0 |
| NEWS2 | Oxygen liters | 0 |
| NEWS2 for COVID | Estimated Glomerular Filtration Rate | 0 |
| CURB-65 | Confusion | 0 |

**Supp Table 6: imputed values for the missing or partially-missing variables of the severity/mortality scores for COVID-19 patients.** We used 0 for imputation as the missing variables are included in linear models only and the constant value does not impact AUC.
