## Supplementary material about segmentation for "Integration of clinical characteristics, lab tests and a deep learning CT scan analysis to predict severity of hospitalized COVID-19 patients"

### Supplementary material about segmentation for the paper “Integration of clinical characteristics, lab tests and a deep learning CT scan analysis to predict severity of hospitalized COVID-19 patients.”

**Annotation scenario of CT scans by radiologists**

Two radiologists (4 and 9 years of experience) examined and annotated 307 anonymized chest scans independently and without access to the patient's clinic or COVID-19 PCR results. All CT images were viewed with lung window parameters (width, 1500 HU; level, -550 HU) using the SPYD software developed by Owkin. Regions of interest were annotated by the radiologists in four distinct classes : healthy pulmonary parenchyma, ground glass opacity, consolidation, crazy-paving. The presence of organomegaly was also notified when present, as a binary class. When multiple CT images were available for a single patient, the image to analyze was selected using the SPYD software. One AI and imaging PhD student also provided full 3D annotation of the four classes on 22 anonymized chest scans using the 3D Slicer software.

**Method to segment CT-scans**

The model used to perform segmentation and compute the AI-*segment* score was based on 3 segmentation networks: 3D Resnet50(Hara, Kataoka, and Satoh 2017) , 2.5D U-Net, and 2D U-Net (Ronneberger, Fischer, and Brox 2015). U-Net consists of convolution, max pooling, ReLU activations, concatenation and up-sampling layers with sections: contraction, bottleneck, and expansion (Supp Fig segmentation 1). ResNet contains convolutions, max pooling, batch normalization, and ReLU layers that are grouped in multiple bottleneck blocks. All models were trained on CT scans provided by Kremlin-Bicêtre (KB) and evaluated on annotated CT scans from Institut Gustave Roussy (IGR). The dataset was divided into two categories: Fully Annotated Scans (FAS) composed of 22 scans (8 from KB and 14 from IGR) and Partially Annotated Scans (PAS) composed of 307 scans (176 from KB and 131 from IGR). PAS contains a total of 7,374 annotated slices and 24,476,521 annotated pixels, i.e. 24 slices per PAS and 3,319 pixels annotated per slice on average.

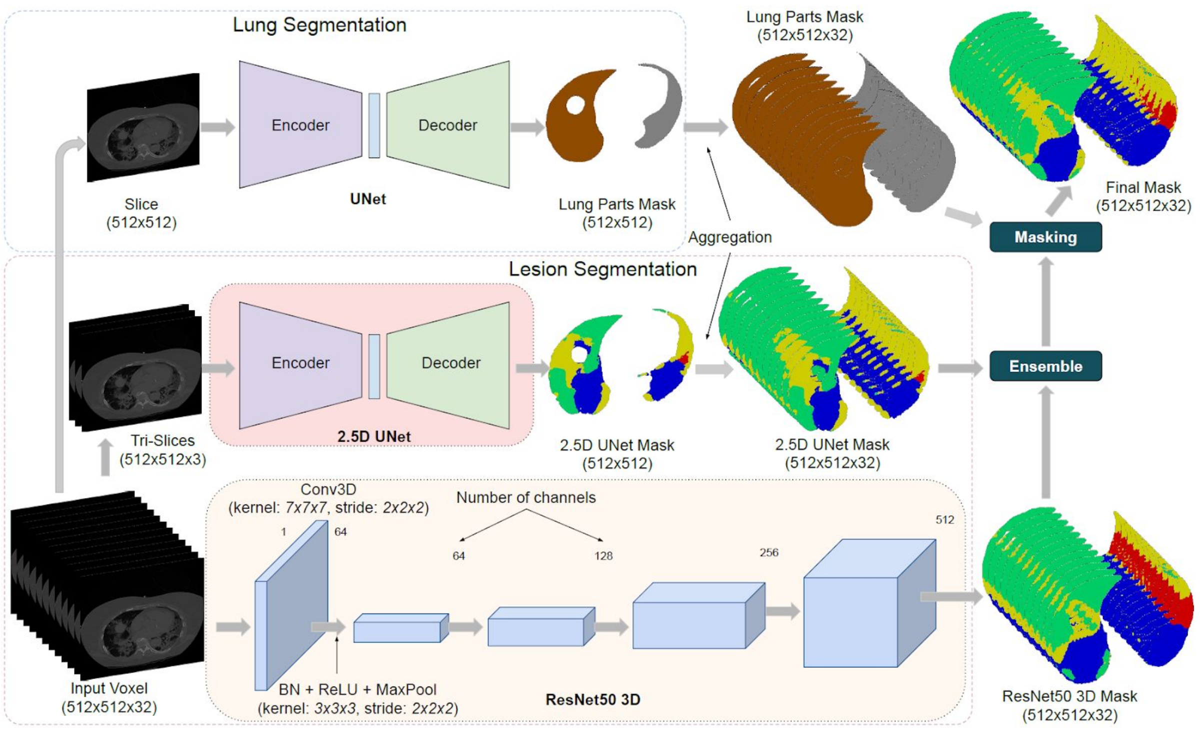


**Supp Fig segmentation 1: architecture of the segmentation model-** Proposed pipeline to generate lesion volumetry estimates from patient CT scans employing ensemble of segmentation networks. Normalized patient scans are provided to our trained 2.5D U-Net and 3D ResNet50. The masks predicted from both models are then merged by arithmetic mean. In parallel, we segment left-right lungs from the patient scans using a dedicated U-Net. Finally, the left-right lung mask is used to mask-out lesions in left and right lungs from the ensemble output. This pipeline utilizes the complementary features learned by a weak model (2.5D U-Net) and a strong one (3D ResNet50).

2D U-Net was trained for left/right lung segmentation and 3D ResNet and 2.5D U-Net were used for lesion segmentation. 3D ResNet50 was trained on 8 KB FAS (i.e. 3,704 slices). Inputs for the 3D ResNet consist of a height and a width of 128, and a depth of 32. We initialized the 3D ResNet with pretrained weights (Chen, Ma, and Zheng 2019). We then trained the network with Stochastic Gradient Descent for parameter optimization and an initial learning rate of 0.1 with a decay factor of 0.1 every 20 epochs. The network was trained for a total of 100 epochs. For the 2.5D U-Net, we first pretrained the network on a left-right lung segmentation task using the LCTCS dataset (Yang et al. 2018). The network was then trained on the KB dataset using Adam optimization algorithm with a learning rate, weight decay, gradient clipping and learning rate decay parameters of 1e-3, 1e-8, 1e-1, and 0.1 (applied at epochs 90 and 150) for 300 epochs. While the validation set remains the same as when evaluating the 3D resnet50 model, 176 KB PAS scans were added to the 8 KB FAS, in the training set. PAS were only added to the 2.5D U-Net training set due to the incompleteness of the annotated volume in the scans which would not satisfy the volumetric requirements of the 3D ResNet50 input. Finally, for the left/right lung segmentation, the 2D U-Net was trained on the 8 KB FAS. Similarly to 2.5D U-Net, Adam optimization algorithm was used with a learning rate, weight decay, gradient clipping, learning rate decay, and number of epochs of 1e-3, 1e-8, 1e-1, 0.1 (applied at epoch 70), and 104. Both 2.5D U-Net and 2D U-Net used affine transformation and contrast change for data augmentation while 3D ResNet50 used affine transformation, contrast change, thin plate splines, and flipping. 3D ResNet and 2.5D U-Net are trained through the minimization of a cross entropy loss and 2D U-Net minimized a binary cross entropy loss. All training was performed on NVIDIA Tesla V100 GPUs using Pytorch as a coding framework. During the validation phase, ensemble inference(Baldeon Calisto and Lai-Yuen 2020) was performed on all available scans by averaging lesion masks, which were predicted from the 3D ResNet and 2.5D U-Net models, using arithmetic mean.

We evaluated the segmentation model on three distinct aspects. First, we evaluated its ability to perform accurate segmentation. To this aim, we computed F1 scores for the PAS (partially annotated scans) and FAS (fully annotated scans), of the IGR test set, when discriminating lesions versus sane areas inside the lung. Micro-averaging was used to limit the effect of class imbalance for the three different lesion types. We also reported the accuracy to discriminate background versus lung regions using FAS where background regions outside of the lung were annotated. Second, we evaluated its ability to estimate the proportion of each lesion type per scan. To this aim, we computed the median, minimum and maximum of the absolute value of the difference between the ground truth percentage of each lesion type obtained from radiologists' annotations and the estimated ones, on the 14 available FAS of the IGR dataset. Third, we evaluated to what extent the segmentation model reproduces the analysis reported by radiologists. To this aim, we first compared the binary decision 'presence or absence of a lesion type' of the network to the radiologist report considered as ground truth. A lesion type was detected by the segmentation model when its estimated volumetry, averaged over both lungs, was above a certain threshold. The difference was then evaluated in terms of detection accuracy and F1 score, for two threshold values, using all scans of the IGR dataset (Supp Table segmentation 1). Then, we compared disease extent as evaluated by radiologists to the one predicted by the neural network (Supp Fig 3).

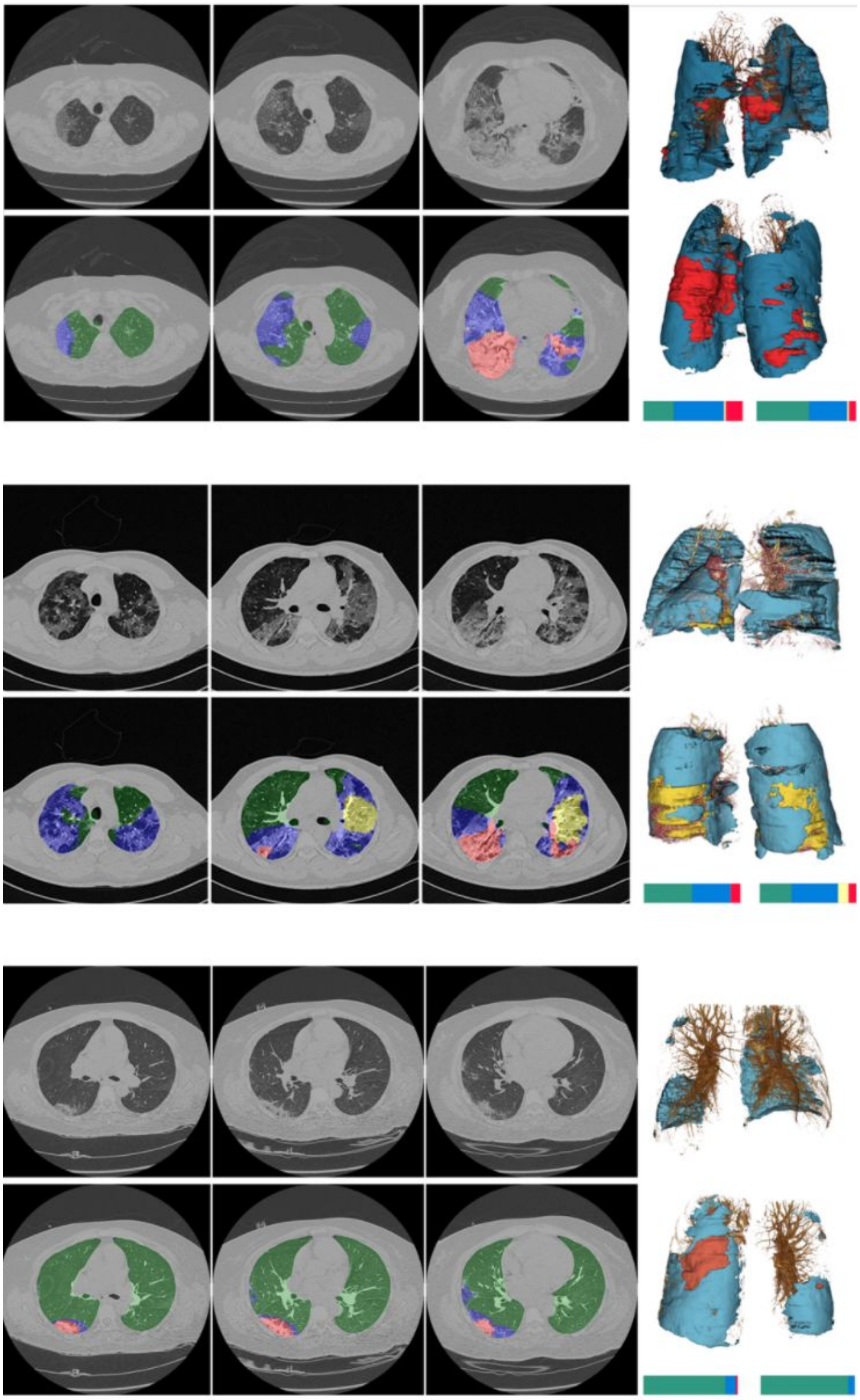

**Supp Fig segmentation 2**: **Axial chest CT scans and segmentation results** COVID-19 radiology patterns, as provided by the neural network model for segmentation, for 3 patients with COVID-19. Green/transparent: sane lung; blue: GGO; yellow : crazy paving; red: consolidation. (Top) Patient with diffuse distribution, and multiple large regions of subpleural GGO with consolidation to the right and left lower lobe. Estimated disease extent by AI: 69%/47% (right/left). Radiologist report: critical stage of COVID-19 (stage 5). (Middle) Patient with diffuse distribution and multiple large regions of subpleural GGO with superimposed intralobular and interlobular septal thickening (crazy paving). Estimated disease extent by AI: 51%/68% (right/left). Radiologist report: severe stage of COVID-19 (stage 4). (Bottom) Patient with minimal impairment, and multiple small regions of subpleural GGO with consolidation to the right lower lobe. Estimated disease extent 13%/7% (left/right). Radiologist report: moderate stage of COVID-19 (stage 2).

| | GGO | Crazy paving | Consolidation |
|---|---|---|---|
| Accuracy (1% thresh.) | 0.7951 | 0.7684 | 0.6167 |
| F1 Score (1% thresh.) | 0.8848 | 0.6452 | 0.7473 |
| Accuracy (2% thresh.) | 0.7876 | 0.7692 | 0.6667 |
| F1 Score (2% thresh.) | 0.8800 | 0.6182 | 0.7848 |

**Supp Table segmentation 1: Detection accuracy and F1 scores of the segmentation model when considering the radiologist report as ground truth.** The binary decision used to compute the score is "presence or not of a lesion type". Accuracy and F1 score are averaged over the IGR validation set. We compared, for each patient of the IGR validation set, detection obtained using AI-*segment* to the information provided in the standardized radiologist report. When using the neural network, a lesion type is considered as present when its relative volume w.r.t. the full volume of both lung, is above a certain threshold indicated into parenthesis in the 1st column of the table.

| **Variable** | **Center** | **Odds ratio** (95% lower - 95% upper) | **P-value** | **P-value Stouffer** |
|---|---|---|---|---|
| GGO AI | KB | 0.61 (0.51,0.73) | 3.57e-08 | 1.37e-08 |
| GGO AI | IGR | 0.77 (0.54,1.10) | 0.15 | |
| Crazy Paving AI | KB | 1.60 (1.29,1.99) | 1.74e-05 | 7.10e-06 |
| Crazy Paving AI | IGR | 1.31 (0.92,1.87) | 0.13 | |
| Consolidation AI | KB | 1.51 (1.27,1.79) | 2.85e-06 | 1.32e-06 |
| Consolidation AI | IGR | 1.27 (0.89,1.82) | 0.19 | |
| Disease extent AI | KB | 2.15 (1.77,2.60) | 7.90e-15 | 1.92e-16 |
| Disease extent AI | IGR | 1.90 (1.30,2.79) | 9e-4 | |

**Supp Table segmentation 2**: **Association between severity and amount of lesions inferred by the segmentation model.** For disease extent, we consider the proportion of lung volume. For the other three variables (GGO, consolidation, crazy paving), we normalize them by disease extent so that each variable measures the proportion of the corresponding lesion.

***Segmentation results for CT-scans of hospitalized COVID-19 patients***

The segmentation model provides automatic quantification of the volume of lesions, expressed as a percentage of the full lung volume (see Supp Fig segmentation 2). These patterns included the three distinguishable features that appear as disease severity progresses: ground glass opacity (GGO), crazy paving, and finally consolidation. The model was trained on 184 patients from KB hospital (8 fully annotated scans, 176 partially annotated ones) and evaluated on 145 patients from IGR hospital (14 fully annotated scans and 131 partially annotated ones). To evaluate the segmentation network, we first compared its performance to that of radiologists manual annotation. The segmentation network discriminated lung regions from regions outside of the lung with an accuracy of 99.9% when evaluated on the fully annotated scans. Within the lung, the model's ability to discriminate between lesions and healthy areas had F1 values of 0.85 and 0.98 on partially and fully annotated scans. In the fully annotated scans, the predicted volumes of each lesion type had relative errors (median [min-max]) of 3.77% [0.054%-14%] for GGO, 0.96% [0.058%-4.4%] for consolidation, and 5.92% [0.41%-13%] for sane lung (no crazy paving was present in these scans). We next compared the segmentation network to the information contained in the radiology reports. The F1 score measuring the ability of the network to detect the presence of a lesion type per patient, was of 0.88 for GGO, 0.65 for crazy paving, and 0.75 for consolidation (Supp Table segmentation 1). Correlation between quantification of the proportion of lesions with the network and the radiologist evaluation was of 0.56 (Supp Fig 3). Inspection of visual results were also consistent with radiologist observations (see Supp Fig segmentation 2 for three representative cases). We lastly evaluated to what extent the segmentation network provided biomarkers of future severity (Supp Table 2 segmentation). We found that severity was significantly associated to GGO extent (OR KB = 0.61 (0.51,0.73), OR IGR = 0.77 (0.54,1.10), $P_{Stouffer}$ = 1.37e-08), crazy paving extent (OR KB = 1.60 (1.29-1.99), OR IGR = 1.31 (0.92,1.87), $P_{Stouffer}$ = 7.10e-06), consolidation extent (OR KB = 1.51 (1.27,1.79), OR IGR = 1.27 (0.89,1.82), $P_{Stouffer}$ = 1.32e-06) as well as total disease extent (OR KB = 2.15 (1.77,2.60), OR IGR = 1.90 (1.30,2.79), $P_{Stouffer}$ = 1.92e-16) (accounting for multiple testing).
